# Caregiver Perspectives of Pre-Transplant Evaluation in Children

**DOI:** 10.1101/2021.06.04.21258216

**Authors:** Eloise C. Salmon, Laura G. Barr, Douglas L. Hill, Judy A. Shea, Sandra Amaral

**Affiliations:** University of Michigan; University of Pennsylvania; Children’s Hospital of Philadelphia; Perelman School of Medicine, University of Pennsylvania

**Keywords:** transplant, pediatric, evaluation, patient experience

## Abstract

Pre-transplant evaluation is mandated by Centers for Medicare and Medicaid Services, but there is wide institutional variation in implementation, and the family experience of the process is incompletely understood. Current literature largely focuses on adult transplant recipients. This qualitative study begins to fill the knowledge gap about family experience of the pre-transplant evaluation for children through interviews with caregivers at a large pediatric transplant center. Prominent themes heard from caregivers include (1) the pre-transplant evaluation is overwhelming and emotional, (2) prior experiences and background knowledge frame the evaluation experience, and (3) frustration with communication among teams is common. These findings are relevant to efforts by transplant centers to optimize information delivery, minimize concrete barriers, and address healthcare systems issues.

## Introduction

For children with advanced chronic kidney disease (CKD) requiring renal replacement therapy, kidney transplant is superior to dialysis, conferring substantially lower risk for morbidity and mortality [1]. Importantly, a child and her family must undergo a comprehensive, interdisciplinary pre-transplant evaluation before scheduling a living donor kidney transplant or qualifying for placement on a deceased donor registry [2]. While this evaluation is mandated, the process is not standardized across centers, and caregiver perspectives of the evaluation are poorly understood.

Prior studies in adults suggest both individual- and systems-level factors play a role in subjective experience of the evaluation as well as time to completion of evaluation [3–8]. Some previously reported individual factors, such as transportation and childcare needs presenting challenges to attending evaluation appointments, also would be relevant for families at a pediatric center. Others, such as fear of comorbid conditions like diabetes and cardiovascular disease making transplant unattainable and limiting motivation for evaluation, would be far less common. System-level factors, particularly communication between transplant center and patient/family, would be universally applicable.

At our center, pre-transplant evaluation presently is initiated by referral letter to the transplant coordinator from a patient’s primary nephrologist. From there, the coordinator contacts the family and schedules two dates, generally non-adjacent, for the family to spend 5-6 hours in-center. Over those two days, appointments typically include the transplant coordinator, transplant nephrologist, transplant surgeon, psychosocial team, financial planner, pharmacist, dietician, infectious disease, and anesthesia. Labs, echocardiography, outstanding immunizations, dental clearance, and ophthalmology, as well as any of the aforementioned appointments unable to be consolidated, are scheduled separately.

Our qualitative study aimed to describe caregiver experience with pre-transplant evaluation for children as well as identify barriers and facilitators to completion of the evaluation for kidney transplant candidates at a large pediatric center in the United States. In particular, we wanted to shed light on how families felt during the evaluation, how the evaluation compared to their expectations, how useful center-specific tools were during the evaluation, and how completely our center answered questions during the evaluation. Additionally, we sought to understand how certain demographic variables, including income and distance from the transplant center, may relate to time to completion of evaluation.

## Materials and Methods

### Study Design and Sample

We conducted a single-center, qualitative study using semi-structured interviews with caregivers of children referred for kidney transplant at our center between July 2017 and December 2018. We excluded caregivers of children who had relocated, were referred as part of multi-organ transplant, or were deceased. Interviews took place between July 2019 and February 2020 and were transcribed verbatim.

The Children’s Hospital of Philadelphia Institutional Review Board approved the study as exempt, but, prior to participation, all caregivers provided written consent for audio recording of the interview as well as permission for the research team to access their child’s medical record. We used a convenience sampling strategy, recruiting caregivers when they were at our center for other appointments.

### Interview Guide

The interview guide (see Supplemental Materials) included both open- and closed-ended questions to collect perspectives on the areas of interest described under Introduction Questions were vetted at institutional research-in-progress meetings and piloted for understandability with families who did not meet eligibility criteria based on referral date.

### Data Collection

Two study authors (E.S. or L.B., neither members of center transplant committee) consented eligible caregivers face-to-face and interviewed them either in-person or by phone, per participant preference. At time of interview, some children already had received their transplant, others were on chronic dialysis or continued with CKD care. Child and family demographic data were collected with an optional electronic questionnaire following the interview as well from the child’s medical chart.

### Analysis

Data analysis commenced at the start of the first interview and was ongoing throughout data collection. We used an emergent, iterative approach where we refined our questioning based on analysis of prior interviews. We continued interviews until our analysis reached thematic saturation with no new themes emerging. Data were analyzed using a consensus approach to inform theme analysis between the two interviewers as we looked for similarities and differences. When differences in interpretation arose, we pursued agreement through discussion. NVivo was used to code transcribed interviews.

## Results

### Participant Characteristics

Of the 56 children referred for kidney transplant between July 2017 and December 2018, 48 (86%) met eligibility criteria for recruitment of their caregivers. Caregivers of twenty-two children signed consents; nineteen completed interviews, fifteen of whom also submitted the written questionnaire. See Table 1 for participant characteristics. The majority of participants were white with household income less than $120,000 and lived less than 100 miles from our center.

**Table 1.**
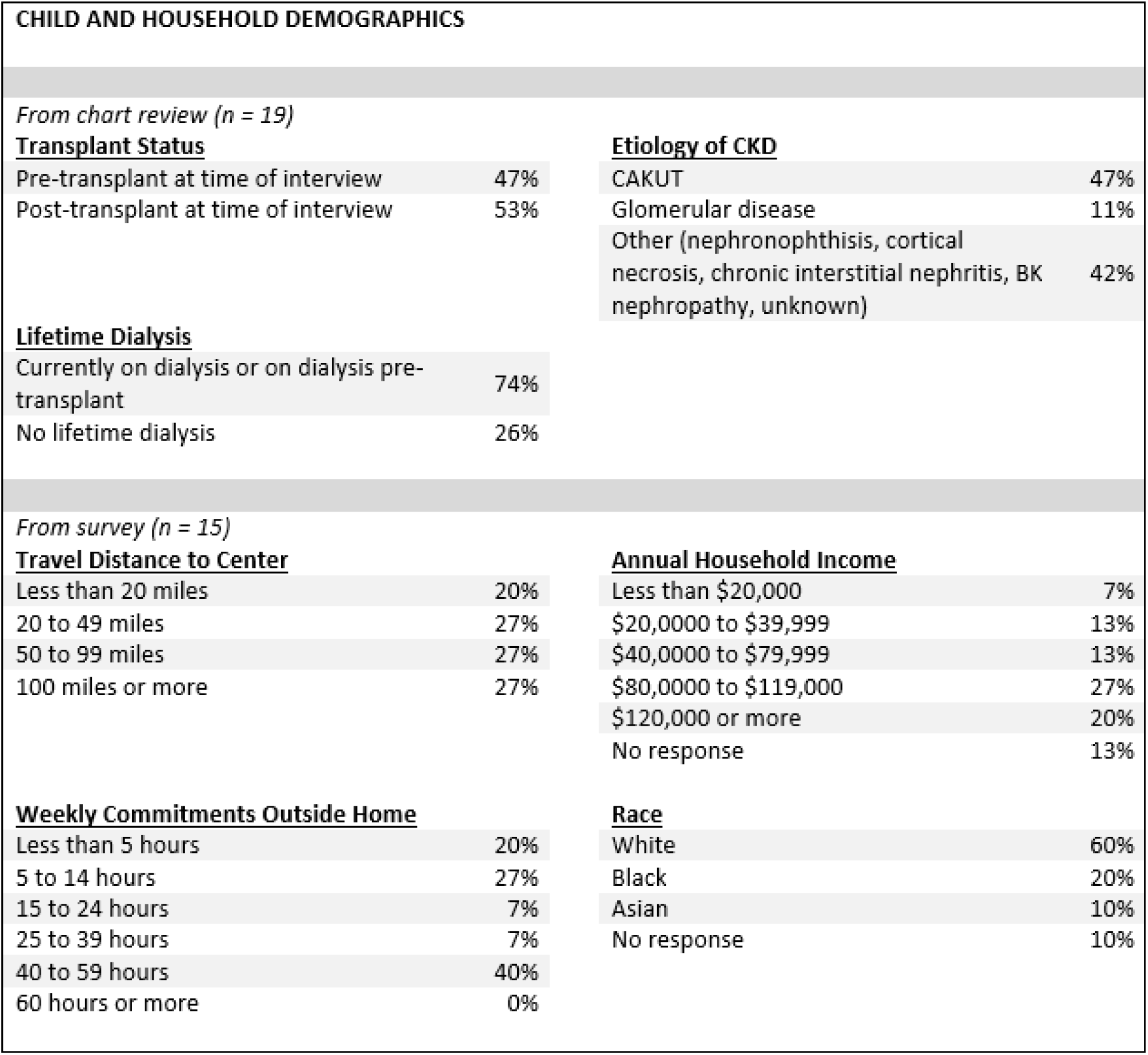
Participant characteristics

### Themes

Prominent themes from the interviews included (1) description of the evaluation as overwhelming and evoking a wide range of emotions, (2) prior experiences and background knowledge as influential, and (3) frustration with communication among teams. Table 2 highlights representative quotations from caregivers on these themes.

**Table 2.**
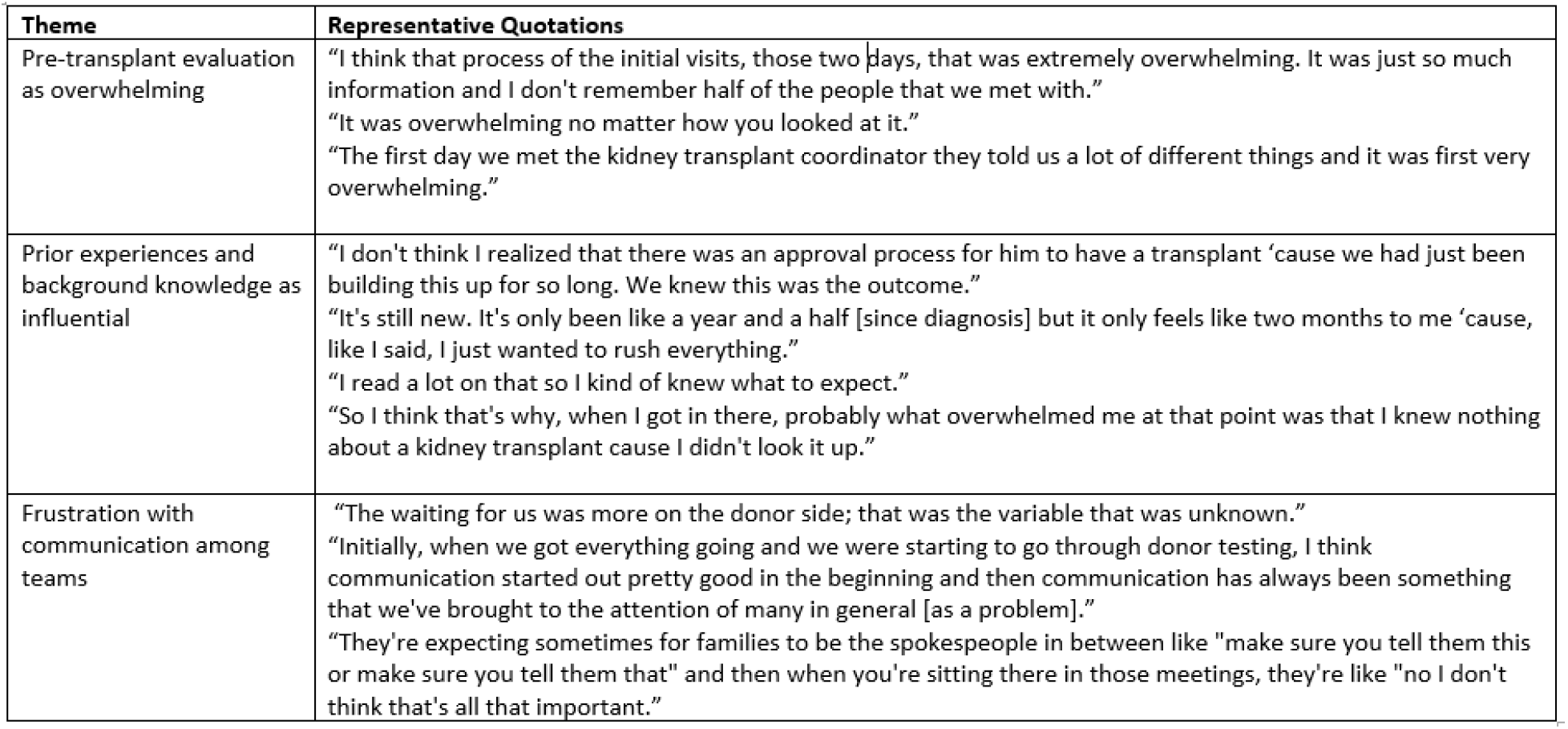
Representative quotations from caregivers according to theme.

| Theme | Representative Quotations |
| --- | --- |
| Pre-transplant evaluation as overwhelming | "I think that process of the initial visits, those two days, that was extremely overwhelming. It was just so much information and I don't remember half of the people that we met with."<br>"It was overwhelming no matter how you looked at it."<br>"The first day we met the kidney transplant coordinator they told us a lot of different things and it was first very overwhelming." |
| Prior experiences and background knowledge as influential | "I don't think I realized that there was an approval process for him to have a transplant 'cause we had just been building this up for so long. We knew this was the outcome."<br>"It's still new. It's only been like a year and a half [since diagnosis] but it only feels like two months to me 'cause, like I said, I just wanted to rush everything."<br>"I read a lot on that so I kind of knew what to expect."<br>"So I think that's why, when I got in there, probably what overwhelmed me at that point was that I knew nothing about a kidney transplant cause I didn't look it up." |
| Frustration with communication among teams | "The waiting for us was more on the donor side; that was the variable that was unknown."<br>"Initially, when we got everything going and we were starting to go through donor testing, I think communication started out pretty good in the beginning and then communication has always been something that we've brought to the attention of many in general [as a problem]."<br>"They're expecting sometimes for families to be the spokespeople in between like "make sure you tell them this or make sure you tell them that" and then when you're sitting there in those meetings, they're like "no I don't think that's all that important." |

### Caregiver Experience of the Pre-Transplant Evaluation as Overwhelming and Evoking a Wide Range of Emotions

Nearly every caregiver used “overwhelm” at some point during the interview, underscoring that families do not take the evaluation lightly. The interview guide also asked caregivers to recall any emotions from during their child’s evaluation. Reponses included a wide range of both positive and negative emotions, with many caregivers experiencing multiple emotions over the course of the evaluation, or even simultaneously. Descriptors with more negative emotional associations included worry, fear, uncertainty, frustrations, depression, “roller coaster”, exhaustion, tears, denial, shock, and anxiety; in contrast, “stoked”, comfort, calm, happiness, confidence, positive thinking, and hope reflected more positive emotions. Caregivers expressed that while team members involved in the evaluation acknowledged the heightened emotions often present in the steps toward transplant, efforts to normalize caregivers’ range of feelings would be appreciated.

### Prior Experience and Background Knowledge as Influential

The range of life paths by which children arrive at referral for kidney transplant quickly emerged as having an important influence on caregiver experience of the evaluation. Some caregivers had been preparing themselves for a child’s transplant since the prenatal period; others had received a diagnosis of kidney problems in their previously healthy child less than a year prior to evaluation. Caregivers in the first group were more likely to (1) point to the evaluation as the moment when it “really sunk in” that a team of transplant-focused physicians, and not their current primary nephrologist, would follow their child post-transplant and (2) wonder if some evaluation requirements (e.g. vaccines) could have happened earlier in their child’s CKD care. In contrast, those in the second group tended to describe the evaluation as part of the same life event as diagnosis. Notably, a long CKD history did not necessarily translate into concrete knowledge about specifics of transplantation before the evaluation. Caregivers who had read previously about kidney transplant (regardless of their child’s time with CKD) felt better able to absorb information during the evaluation. Notably, some caregivers who had managed complex CKD regimens for years acknowledged that they did not appreciate how much new information transplant would entail until the evaluation.

### Frustrations with Communication among Teams

While gathering input from relevant subspecialists and living donor evaluation are separate from the core elements of a child’s evaluation, caregivers in these situations expressed frustration that the relevant teams did not interface with each other more seamlessly and communicate group decisions to the family more effectively. Families felt these frustrations both within the pediatric center as well as between the pediatric center and the living donor evaluation center. For example, if the initial days of the child’s evaluation revealed that a pediatric subspecialist outside the core transplant team would need to weigh in on readiness for transplant, caregivers at times felt like intermediaries conveying information among providers, unclear whether the transplant team or the subspecialist needed to make the next decision. Similarly, if the caregiver hoped to be a living donor, it was perceived to be difficult to ascertain the status of the donor evaluation.

### Barriers and Facilitators to Completion of Evaluation from Written Questionnaire Data

The interview guide contained items to assess whether specific institutional features might present unrecognized barriers to completion of evaluation, including asking caregivers whether any particular element was difficult to complete, or if they felt stuck or disrespected at any point. No element was identified recurrently, and no caregiver recalled an instance of disrespect.

While no caregiver pointed to other demands on time as a barrier to completion of the evaluation during the interview, data from the categorical questionnaire suggest more hours each week of commitments outside the home may be associated with longer time to completion of evaluation. Of the seven caregivers whose children had a complete evaluation within 3 months, five had less than 15 hours of outside commitments per week (one more than 15 hours, one no response). In contrast, for those with time to completion of more than three months, most had weekly commitments exceeding 15 hours. Similarly, caregivers whose child completed the evaluation in less than three months had one or two children in the household and lived less than 50 miles from our center whereas those caregivers whose child took more than three months to complete the evaluation often had three or more children in the household and lived 50 miles or more from our center. No similar pattern was seen for either income or education.

As for possible facilitators, the interview guide solicited feedback on two tools available to caregivers in navigating the evaluation process: a printed road map (see Supplemental Materials) and the patient portal of the electronic medical record (EMR). The road map received neutral to positive reviews, with some families finding it especially helpful when someone from the transplant team wrote in appointment dates. One caregiver with a background in communications and graphic design had several ideas about how to improve the roadmap, including capturing the evaluation as part of the continuum from CKD care through post-transplant. As for the EMR, some caregivers used it regularly during the evaluation to track appointments.

### Suggestions for Improvement

Beyond the ideas captured in the sections above, caregivers offered additional suggestions for improvement when asked what else we should know about their evaluation experience in looking ahead to the care of future families. Some concrete ways to improve evaluation process and content included:

- A book with photos not only identifying the name and title of everyone on the transplant center team, but also describing for what reasons the caregiver should contact them and their role in pre- and post-transplant processes
- A scheduled phone call some weeks after the initial evaluation visit devoted entirely to soliciting follow up questions
- Inclusion of information in the evaluation about what and when to share with a child after receiving a call for a possible organ offer (“he [the child] was absolutely devastated when he didn’t get it”)
- Increased emphasis on range of post-operative experiences to feel more prepared for a hospital stay either shorter or longer than quoted averages at our center (5 days for an adolescent, 7-10 days for a younger child)
- Identifying support groups where caregivers can hear about experiences of other families with children who are waiting for or have received a kidney transplant, including center-sponsored events where caregivers could “brainstorm” together

## Discussion

The interviews for this study made clear that the pre-transplant evaluation is a significant life event for caregivers – overwhelming at times, full of heightened emotions, colored by past experiences, and containing a large volume of information. This raises the question of whether a decompressed core evaluation over more days would be preferable, but no caregiver expressed an interest in this prospect as it likely meant more travel and more missed days of school/work. Importantly, several of the suggestions for improvement above, including a book with team member photos and a scheduled follow up phone call, are not particularly resource intensive and may go a long way in helping caregivers process content and direct questions.

The apparent consensus among caregivers on keeping in-center days to a minimum aligns well with the observed association between hours of weekly commitments, number of children in household, and travel distance with time to completion. With these data in mind, greater use of telehealth is an intriguing option. Social distancing guidelines during the COVID19 pandemic dramatically increased uptake of this technology at our center; going forward, it may be a useful tool in conducting certain evaluation elements for maximal convenience of both providers and families. For example, slides covering basic concepts of transplant could be presented effectively via a remote meeting platform, and elements like pharmacy and financial clearances may become even more robust by creating a setting where families have easier access to items like current pillboxes and insurance records.

To optimize caregiver understanding and comfort with the evaluation, there may be a role for distributing selected educational materials in advance. Understanding whether it would be most useful for families to receive materials as printed/electronic brochures (such as “What Every Parent Needs to Know: A guide when your child needs a transplant”, UNOS 2018), online videos/webinars, or in some other format is an area rich for future study, innovation and quality improvement across centers. Future work also could explore when and how basic principles of and requirements for transplant are discussed by primary nephrologists during routine care earlier in a child’s CKD course. Many families form an unusually close relationship with the primary nephrologist, in some cases since the patient’s infancy (or even prenatal counseling). Transplant teams can build on this relationship to promote a positive evaluation experience for families.

Caregivers repeatedly underscored that clear communication is key, whether in-person or via other means. The interface between the evaluation of the child and the evaluation of living donors was a source of frustration for many caregivers. To families, the evaluations feel like part of a single life event, and the separation between different medical silos seem senseless to them. There may be value in redoubling efforts to highlight the ethical reasons for separate evaluations as well as the logistical realities (e.g. unlike for their children, caregivers cannot just call an adult center for the test results of their spouse.) Designating a team member as the point person for questions or concerns about living donor evaluation also may improve caregivers’ experience of this interface. Similarly, processes for obtaining and documenting considerations and recommendations from other subspecialists should be clarified, so caregivers never feel the burden to gather this information is on them.

In addition to guiding local changes, these findings have broader policy implications. Most importantly, any effort to move toward a hybrid clinic/telehealth evaluation for increased family-centeredness necessitates clarification of billing regulations. As a complementary step, the observed association between time to completion of evaluation and weekly commitments/number of children further supports current advocacy efforts to protect time off work and cover childcare expenses during the evaluation. Similarly, reimbursement for personnel to serve as liaisons also could expedite completion of evaluation requirements and improve family experience. In limited-resource settings where such program expansion may not be viable, support and guidance from CMS and UNOS on best practices for (1) delivering information to optimize informed consent and (2) promoting effective communication across a health system also could have benefit at less expense.

### Limitations

This study has several limitations. First, caregivers were recruited from a single center so their experiences may not generalize nationally or internationally, but most themes seem relevant to workflows at other centers. Second, we interviewed caregivers of less than half of the children referred for kidney transplant between July 2017 and December 2018. While the content of later interviews supported good thematic saturation, it is possible that important perspectives are not represented here. For example, in conceiving this project we were particularly motivated by children who do not complete all elements of the pre-transplant evaluation in one year and end up starting much of it over. Especially given the possible associations seen here between time to completion of evaluation and demands from outside commitments, other children, and travel, future qualitative studies should focus on the caregivers of this subset of children to improve our knowledge of their experience of the evaluation. Finally, understanding how the child experiences the evaluation also is critical in increasing family centeredness of the evaluation process and was not addressed here.

## Conclusion

This qualitative study is the first to our knowledge to focus on the caregiver experience of the pediatric kidney transplant evaluation. Broad themes around volume of information and team communication resonated with prior work in the adult literature, but the experience of a caregiver as a potential living donor as well as the natural history of CAKUT and the resulting relationship between a caregiver, child, and their primary nephrologist are more specific to pediatric transplant. These data will inform efforts for ongoing improvement in the pre-transplant evaluation process by highlighting the importance of (1) acknowledging the scope of content and continually reevaluating accessibility of delivery (2) recognizing the influence of prior experience and tailoring process elements accordingly for increased family-centeredness and (3) making concerted efforts to define roles and set expectations, especially when multiple teams or institutions are involved in care. The findings not only are relevant to transplant centers, but also to broader commitments across the nephrology community to optimizing delivery of information about complex health care topics.

## Data Availability

The data generated during the current study are available from the corresponding author on reasonable request.

## Funding

ES was supported by a pilot grant from the Center for Pediatric Clinical Effectiveness at the Children’s Hospital of Philadelphia and a T32 training grant supervised by Dr. Lawrence Holzman (DK007006-46). SA is supported by R01 DK120886, R01 DK110749 and R01 HD091185.

## Conflicts of interest/Competing interests

No authors have conflicts to disclose.

## Code availability (software application or custom code)

N/A

## Authors’ contributions

Eloise Salmon, Sandra Amaral, and Judi Shea contributed to the study conception and design. Material preparation and data collection were performed by Eloise Salmon and Laura Barr. All authors contributed to interpretation of data. The first draft of the manuscript was written by Eloise Salmon and all authors commented on previous versions of the manuscript. All authors read and approved the final manuscript.

## Consent to participate

This work was exempt by the Institutional Review Board of the Children’s Hospital of Philadelphia. Participants granted specific permissions for both audio recording of interviews and access to medical record for collection of basic clinical information.

## Consent for publication

Participants made aware of intent to publish prior to their agreement to participate.

## Acknowledgements

Thank you to the CHOP Center for Pediatric Clinical Effectiveness for providing pilot grant funding. ES also is supported by T32 training grant supervised by Dr. Lawrence Holzmann (DK007006-46). SA is supported by R01 DK120886, R01 DK110749 and R01 HD091185.

## Notes

### Competing Interest Statement

The authors have declared no competing interest.

### Funding Statement

ES was supported by a pilot grant from the Center for Pediatric Clinical Effectiveness at the Childrens Hospital of Philadelphia and a T32 training grant supervised by Dr. Lawrence Holzman (DK007006-46). SA is supported by R01 DK120886, R01 DK110749 and R01 HD091185.

### Author Declarations

This work was exempt by the Institutional Review Board of the Childrens Hospital of Philadelphia. Participants granted specific permissions for both audio recording of interviews and access to medical record for collection of basic clinical information.

