## Supplemental Material for "Caregiver Perspectives of Pre-Transplant Evaluation in Children"

INTERVIEW QUESTIONS

1. How would you describe your experience with the pre-transplant evaluation process?
2. How did the overall pre-transplant evaluation process compare to your expectations?
    a. Much easier than expected
    b. Somewhat easier than expected
    c. About the same as expected
    d. Somewhat harder than expected
    e. Much harder than expected
3. How difficult was it to complete each part of the evaluation process? (categorize each as much easier than expected to much harder than expected, or rank relative to each other)
    a. Surgery appointment
    b. Nephrology appointment
    c. Infectious disease appointment
    d. Anesthesia appointment
    e. Dentist appointment
    f. Ophthalmology (eye) appointment
    g. Labwork (blood tests)
    h. Vaccines/immunizations
4. Tell me more about what made these parts easy or difficult.
5. Were there any points in the evaluation process where you felt "stuck"? Tell me more about them.
6. Parents describe experiencing many different emotions during the pre-transplant evaluation. Will you describe to me some of your emotions during the evaluation process?
7. In a process like pre-transplant evaluation that involves multiple parts and many interactions with the health care system, parents sometimes feel disrespected by individuals or by the system. Was there any time during the transplant evaluation that you felt disrespect? Tell me more about it.
8. Was the evaluation roadmap helpful to you during the pre-transplant evaluation process?
    a. Yes, it was very helpful
    b. Yes, it was somewhat helpful
    c. No, it was not helpful
    d. I'm not familiar with the roadmap.
9. Tell me more about what made the roadmap helpful (or unhelpful). How could we improve it?
10. Was myCHOP, the hospital's online patient portal, helpful to you during the pre-transplant evaluation process?
    a. Yes, it was very helpful
    b. Yes, it was somewhat helpful
    c. No, it was not helpful
    d. I do not use myCHOP.
11. Tell me more about what made myCHOP helpful (or unhelpful).
12. How many of your questions about kidney transplant were answered during the pre-transplant evaluation process?
    a. All of my questions were answered
    b. Most of my questions were answered
    c. Some of my questions were answered
    d. Only a few of my questions were answered
    e. None of my questions were answered
13. Anything else our team should know about your experience with the pre-transplant evaluation process? (free response)

Thank you for taking the time to answer these questions. We now would like you to complete a short list of multiple choice questions in writing.

ROADMAP

### Kidney Transplant Process

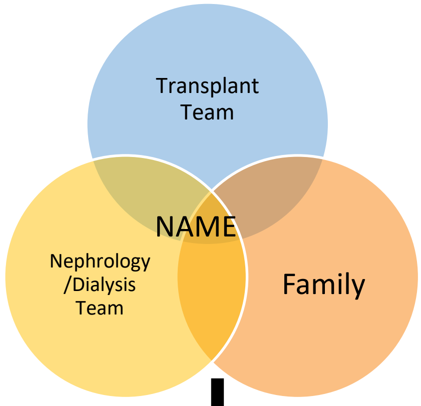


**Eval Start Date**

##### Transplant Coordinator

- Family Meeting
- Transplant Coordinator
- Psychosocial Clearance
- Financial Clearance
- Pharmacy Clearance
- Vaccinations Up to Date – await ID’s recommendations

##### Family

- Transplant Surgeon Consultation Scheduled
- Transplant Coordinator, scheduled
- Transplant Nephrologist, Scheduled
- Infectious Disease (ID) Consultation,
- Nutrition Clearance
- Anesthesia Evaluation
- Dental
- Ophthalmology

##### Labs/Imaging/Testing

- Labs: Blood Type
- Labs: Blood Type
- Labs: HLA testing
- Labs: Viral Studies
- Echocardiogram
- TB screening
- Any additional appointments as needed
